# Development and validation of a knowledge-driven risk calculator for critical illness in COVID-19 patients

**DOI:** 10.1101/2020.06.14.20103754

**Authors:** Amos Cahan, Tamar Gottesman, Michal Tzuchman Katz, Roee Masad, Gal Azulay, Dror Dicker, Aliza Zeidman, Evgeny Berkov, Gal sahaf Levin, Boaz Tadmor, Shaul Lev

## Abstract

Facing the rapidly spreading novel coronavirus disease (COVID-19), evidence to inform decision-making at both the clinical and policy-making level is highly needed. Based on the results of a study by Petrilli et al, we have developed a calculator using patient data at admission to predict the risk of critical illness (intensive care unit admission, use of mechanical ventilation, discharge to hospice, or death).

We report a retrospective validation of the risk calculator on 145 consecutive patients admitted with COVID-19 to a single hospital in Israel. Of the 18 patients with critical illness, 17 were correctly identified by the model(sensitivity: 94.4%, 95% CI, 72.7% to 99.9%; specificity: 81.9%, 95% CI, 74.1% to 88.2%). Of the 127 patients with non-critical illness, 104 were correctly identified. This, despite considerable differences between the original and validation study populations.

Our results show that data from published knowledge can be used to provide reliable, patient level, automated risk assessment, potentially reducing the cognitive burden on physicians and helping policy makers better prepare for future needs.

## INTRODUCTION

Facing the rapidly spreading novel coronavirus disease (COVID-19), evidence to inform decision-making at both the clinical and policy-making level is highly needed [1]. In an impressive work, Petrilli and coworkers [2] have recently reported a multivariable analysis of data collected on 2,729 hospitalized patients with COVID-19 at an academic health system in New York City (NY population), to predict critical illness (defined as a composite of care in the intensive care unit, use of mechanical ventilation, discharge to hospice, or death). Based on this analysis, we have developed a computed calculator for risk stratification of hospitalized COVID-19 patients. Since rates of severe disease and mortality vary widely by country [3], we aimed to validate the risk calculator on a population of COVID-19 patients in Israel (IL population).

## METHODS

### Risk calculator development

We used the odds ratios obtained by Petrilli et al through multivariable regression to develop a risk calculator. These included demographics, past medical history, temperature and oxygen saturation on presentation,and selected labs (the first result of c-reactive protein, d-dimer, ferritin, procalcitonin, and troponin). In their analysis, Petrilli et al found that the risk of developing critical illness was considerably lower for patients admitted later during the study period. This observation was limited to the 6 weeks of the study period and may be the result of unique and local circumstances. Thus, we did not include the week of presentation as a feature in the risk calculator.

### Validation process

We studied patients admitted to the Rabin Medical Center, HaSharon Campus, a teaching medical center at Petach Tikva, Israel. HaSharon Hospital was designated a coronavirus care center, to which COVID-19 patients residing in Central Israel were referred. Included were patients hospitalized starting from March 9, 2020 with confirmed Covid-19, defined as a positive result on real-time reverse transcriptase-polymerase-chain-reaction (RT-PCR) assay of nasopharyngeal or oropharyngeal swab specimens. This was done using a kit by Seegene (Songpa-gu, South Korea). At the time of data extraction, there were no patients still hospitalized who had not reached the composite outcome. Patients’ electronic health records were reviewed by two clinicians (MTK and RM) and relevant demographic data, clinical findings and results of the first laboratory and imaging studies done during the admission were extracted. Based on their state at the time of data extraction, patients were determined to have critical illness (at least one of ICU admission, mechanical ventilation or death; no COVID-19 patients were transferred to hospice care) or non-critical disease. Extracted data was then loaded to the risk calculator and analysis of the relationship between predicted risk and actual outcome was performed.

### Statistical analysis

Analysis was done using R (version 3.6.3)[4]. Differences between the NY and IL populations were assessed using the one sample t-test and one sample Wilcoxon signed rank test. Confidence intervals for the calculator performance measures were computed using the “exact” Clopper-Pearson method.

The study was approved and informed consent waived by the Rabin Medical Center Institutional Review Board (ref. 0339-20-RMC).

## RESULTS

A total of 145 patients were admitted to HaSharon Hospital between March 9, 2020 and May 13, 2020. At the time of analysis,137 had been discharged, none were still hospitalized, and 8 had deceased. Table 1 shows a comparison of the IL and NY populations baseline characteristics. The median age in the IL and NY populations was similar (62 and 63, respectively), and in both, the majority of patients were males (62.1% vs 61.3%). There were no patients of race other than white in the IL population. Fever and oxygen saturation at presentation were significantly higher, on average, in the IL population. Of note, procalcitonin levels were not available for any of the patients, as this test is not routinely performed at HaSharon hospital. Values of other predictors used in the calculator are shown in Table 2. Results of serum troponin, ferritin and d-dimer from the first 48 hours of admission were missing in 31(21.3%), 30(20.7%), and 40(27.6%) of the patients, respectively.

**Table 1:** Characteristics of hospitalized patients.

| **Characteristic** | **NY population** | **IL population** | **p-value** |
| --- | --- | --- | --- |
|  | Hospitalized, N=2,729<br>N (%) or median (IQR) | Hospitalized, N=145<br>N (%) or median (IQR) |  |
| Age | 63 (51-74) | 62(46, 71) | <0.010 |
| Male sex | 1672 (61.3) | 90(62.1) | 0.849 |
| White race | 1089 (39.9) | 145(100) | <0.010 |
| Current smoker | 141 (5.2) | 18(12.4) | <0.010 |
| Obesity | 1081 (39.6) | 33(22.7) | <0.010 |
| Any chronic condition* | 2176 (79.7) | 113(77.9) | 0.609 |
| Hyperlipidemia | 1157 (42.4) | 60(41.3) | 0.859 |
| Hypertension | 1693 (62.0) | 59(40.7) | <0.010 |
| Coronary artery disease | 602 (22.1) | 14(9.6) | <0.010 |
| Heart failure | 349 (12.8) | 1(0) | <0.010 |
| Diabetes | 950 (34.7) | 40(27.6) | 0.058 |
| Asthma or chronic obstructive pulmonary disorder | 453 (16.5) | 14(9.6) | <0.010 |
| Chronic kidney disease | 580 (21.3) | 7(4.8) | <0.010 |
| Cancer | 292 (10.7) | 16(11.0) | 0.90 |
| Temperature at presentation, degrees Celsius | 37.4 (36.9-38.2) | 38.5(38.1, 39.0) | <0.010 |
| Temperature ≥ 38 at presentation | 846 (31.0) | 86(59.3) | <0.010 |
| Oxygen saturation <88% | 422 (15.5) | 6(4.1) | <0.010 |
| Oxygen saturation at presentation | 94 (90-96) | 97(95, 99)** | <0.010 |
| *Not used in the model |  |  |  |
| ** Ambient air |  |  |  |

**Table 2:** Values of first laboratory tests used in the risk calculator^*^.

| **Table 2: Values of first laboratory tests used in the risk calculator\*** |  |  |
| --- | --- | --- |
| **Test** | **Units** | **Median(IQR)** |
| Lymphocytes | 10<sup>3</sup>/uL | 1.0 (0.8, 1.5) |
| Creatinine | mg/dL | 0.88 (0.75, 1.05) |
| C-reactive protein | mg/L | 0.37 (0.09, 1.10) |
| D-dimer | ng/mL | 602.0 (336, 1125.7) |
| Ferritin | ng/mL | 388.1 (161.7, 689.5) |
| Troponin-T | ng/mL | 8.0 (6.0, 14.0) |
| * Procalcitonin testing is not performed at HaSharon hospital |  |  |

Of the 18 patients with critical illness, 17 were correctly identified by the model when a threshold score of 2.2 points was used (sensitivity: 94.4%, 95% CI, 72.7% to 99.9%; specificity: 81.9%, 95% CI, 74.1% to 88.2%). Of the 127 patients with non-critical illness, 104 were correctly identified. The accuracy was 0.83 (95% CI: 0.76-0.89) and the C-statistic, or the area under the ROC (receiver operator curve) was 0.943 (Figure 1).

**Figure 1:**
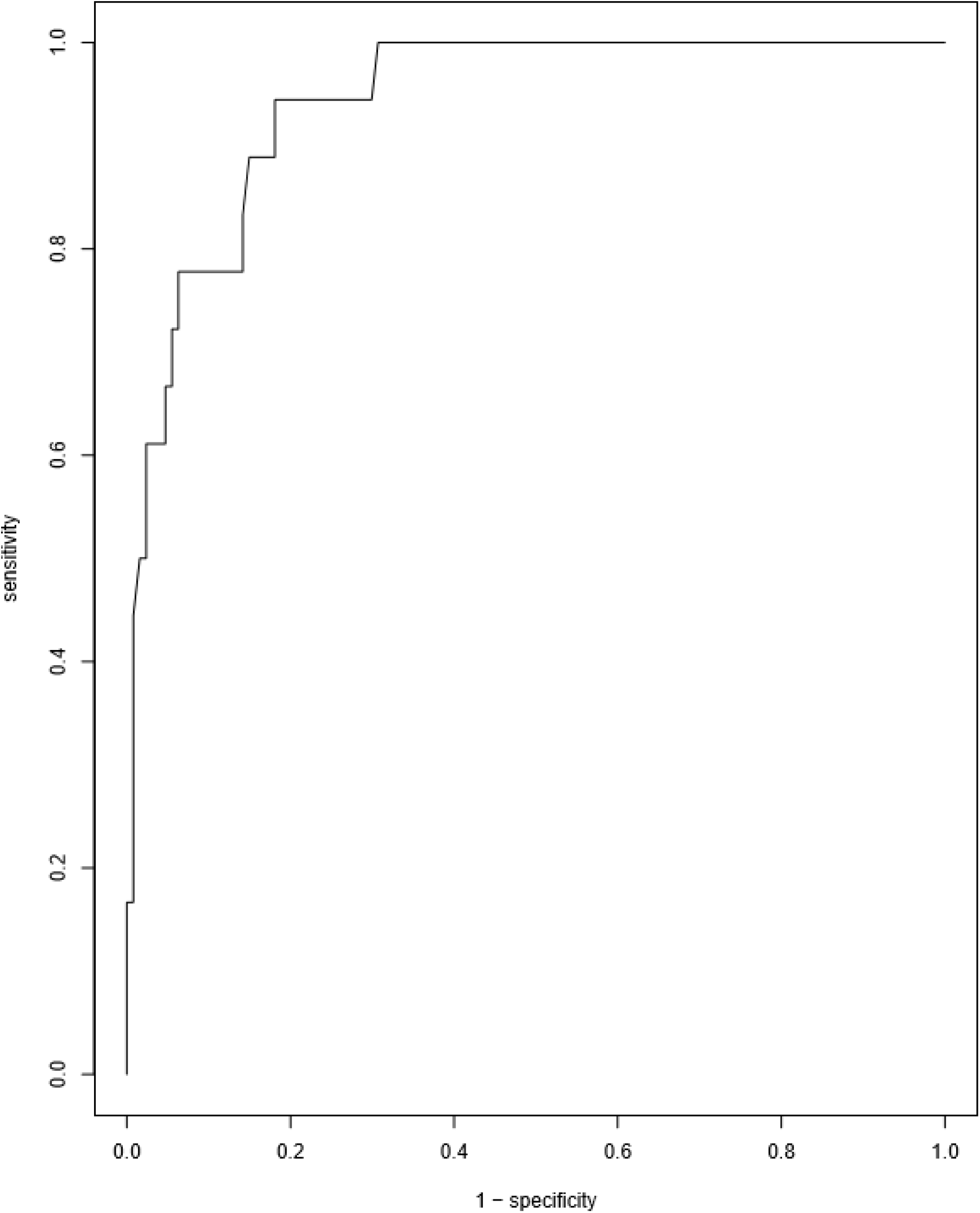
ROC curve of risk calculator performance. AUC (area under the curve): 0.943

## DISCUSSION

We used the results of an analysis performed by Petrilli on a population of COVID-19 in NY to develop a risk calculator for critical illness. We report the successful application of this calculator to make predictions on the outcome of COVID-19 hospitalized patients in Israel. This, despite considerable differences in baseline characteristics between the two populations.

Thousands of papers have been published to date on COVID-19. For clinicians, keeping current on medical literature is challenging in normal times but even more demanding during the COVID-19 pandemic, when their abilities are stretched to the limit. Moreover, given the limited capacity of humans (including physicians) to apply probabilistic reasoning [5], integration of published evidence probably remains mostly at the intuitive level in the minds of clinicians. This state of affairs calls for equipping clinicians with reliable tools to properly evaluate the abundant empirical knowledge, and properly weigh it against their own patients’ data. Our study shows that risk assessment could be done automatically (without active physician involvement), reducing the time and effort required from physicians for risk assessment, and providing important information at scale to policy makers.

Public health measures, such as quarantine and shelter-in-place, are guided by the capacity of the healthcare system, with ICU beds and ventilator availability being the “rate limiting factor”. As severe disease often develops during the second week of illness, there is a reported 12 day average lag between illness onset and ICU admission or the development of acute respiratory distress syndrome (ARDS) [6]. In an effort to avoid overwhelming the healthcare system capacity, further lifting of restrictions on social interactions is thus delayed until the effects of policy changes can be measured. The turnaround time for policy-makers to get feedback on policy changes is therefore around two weeks. Our results provide reason to believe that future critical illness can be reliably predicted at the time of admission.Such predictions may be used in triage to make sure that high risk patients remain where medical care is rapidly available, or to select patients for (investigational) interventions. At the institutional and healthcare system level, predicting the future burden of critical illness could improve allocation of resources (e.g., personnel and supplies) to preempt shortages. At the State and National levels, such predictions could shorten the feedback turnaround time, allowing policy makers to effectively flatten the epidemic curve and avoid breakdown of medical care, while minimizing restrictions on the workforce to curb the financial crisis.

The proposed risk calculator is limited in the sense that it is based on a single study, which, albeit large in scale, includes patients from a single metropolitan area, presenting during a relatively short period of time, in which the local healthcare system was heavily burdened by scores of severely ill patients. Further research will be needed to validate its performance in other patient populations. Another limitation is the lack of information on some laboratory tests which were included in the original analysis, however this could have weakened our results.

## CONCLUSION

We believe that computer-aided risk assessment is a means to put research-derived knowledge to work in the clinical setting. As new research is published, other calculators could be developed. Based on local circumstances, a calculator using predictors from the most relevant study could be used. Moreover, several calculators could be applied in parallel, and calculator voting used to derive potentially more robust predictions.

## Data Availability

Access to the data used in this study is restricted

